# CovidMulti-Net: A Parallel-Dilated Multi Scale Feature Fusion Architecture for the Identification of COVID-19 Cases from Chest X-ray Images

**DOI:** 10.1101/2021.05.19.21257430

**Authors:** Md. Saikat Islam Khan, Anichur Rahman, Md. Razaul Karim, Nasima Islam Bithi, Shahab S. Band, Abdollah Dehzangi, Hamid Alinejad-Rokny

## Abstract

The COVID-19 pandemic is an emerging respiratory infectious disease, having a significant impact on the health and life of many people around the world. Therefore, early identification of COVID-19 patients is the fastest way to restrain the spread of the pandemic. However, as the number of cases grows at an alarming pace, most developing countries are now facing a shortage of medical resources and testing kits. Besides, using testing kits to detect COVID-19 cases is a time-consuming, expensive, and cumbersome procedure. Faced with these obstacles, most physicians, researchers, and engineers have advocated for the advancement of computer-aided deep learning models to assist healthcare professionals in quickly and inexpensively recognize COVID-19 cases from chest X-ray (CXR) images. With this motivation, this paper proposes a “CovidMulti-Net” architecture based on the transfer learning concept to classify COVID-19 cases from normal and other pneumonia cases using three publicly available datasets that include 1341, 1341, and 446 CXR images from healthy samples and 902, 1564, and 1193 CXR images infected with Viral Pneumonia, Bacterial Pneumonia, and COVID-19 diseases. In the proposed framework, features from CXR images are extracted using three well-known pre-trained models, including DenseNet-169, ResNet-50, and VGG-19. The extracted features are then fed into a concatenate layer, making a robust hybrid model. The proposed framework achieved a classification accuracy of 99.4%, 95.2%, and 94.8% for 2-Class, 3-Class, and 4-Class datasets, exceeding all the other state-of-the-art models. These results suggest that the “CovidMulti-Net” framework’s ability to discriminate individuals with COVID-19 infection from healthy ones and provides the opportunity to be used as a diagnostic model in clinics and hospitals. We also made all the materials publicly accessible for the research community at: https://github.com/saikat15010/CovidMulti-Net-Architecture.git.

## Introduction

Computational tools have been extensively utilized to tackle challenging problems in the field of medical science [1]. They have been demonstrated as fast and cost-effective alternative to experimental methods. It is very common to face newer cases of medical complexities over the period of time and numerous experts work seamlessly for days to analyze the problems and find a solution to them. Some disease could be so spreadable that many people lost their lives before our doctors and medical engineers came to a stable outcome. For instance, COVID-19 is the very recent disease that took a lot of lives within just a few months all over the world. The procedure of identifying this viral disease is complex and time consuming. People in Bangladesh (for example) need to wait at least one week to get the report of the COVID-19 test. Apart from this, sample collection process is also problematic and complex. Speeding up the virus can dramatically speed up the treatment and prevention processes. Researchers from all over the world invented different mechanism to detect the virus but still people are facing difficulties.

Currently, chest X-ray (CXR) is a medical imaging technique that helps doctors finding the COVID-19 cases more quickly. As COVID-19 attacks the lungs, the effect could be on the screen of the chest X-ray images. However, it requires expertise and time to test the CXR images manually. Nowadays, Computer-Aided Diagnosis (CAD) system has been designed for detecting COVID-19 cases early without the need for human intervention. In medical imaging technology, the CAD system has improved dramatically due to machine learning (ML) and deep learning (DL) applications. Such techniques lead to achieving high accuracy in terms of detecting COVID-19 cases. However, the prediction performance of this task using classical machine learning methods has remained limited. On the other hands, the deep learning framework has been introduced to address the shortcomings of traditional machine learning techniques by automatically performing the task of feature extraction in a fully end-to-end fashion and learn the input images’ holistic features from low to high level for classification purposes [2]. In addition, by adding transfer learning capabilities, weights of different pre-trained deep learning models which were previously trained on the ImageNet database, can be transferred to perform new relevant classification tasks [3]. Since these models are deeper than the conventional approaches and use more dynamic relations among the alternating layers to detect more complex features, they can achieve significant performance [4].

In this study, we present an automated fast screening of COVID-19 detection system using a “CovidMulti-Net” framework. This framework consists of three CNN pre-trained models including DenseNet-169, ResNet-50, and VGG-19, where each model is used for extracting features from the CXR images. We use a concatenate layer for combining all the extracted features, to build a robust parallel robust model. Finally, we evaluate the performance of our “CovidMulti-Net” framework on three publicly accessible datasets, where we have obtained significant results. The main contributions of this research work are summarized as follows:

- The proposed “CovidMulti-Net” is based on hybrid feature extraction technique that is developed through combining multiple deep CNN pre-trained models.
- The “CovidMulti-Net” framework does not require any segmentation–based feature extraction technique.
- The “CovidMulti-Net” framework does not misclassify any CXR image of the COVID-19.

### Organization

The rest of this article is organized ad follow. The literature overview is presented in Section II. After that, the proposed “CovidMulti-Net” framework is presented in detail, along with data pre-processing and data augmentation strategies in Section III. The performance of the proposed framework is thoroughly discussed in section IV. In Section V, the comparative outcome analysis with future direction is shown. Finally, the conclusion of this work is presented in Section VI.

## Literature Overviews

Recently, studies have proposed different machine learning and deep learning models for detecting COVID-19 diseases. In this section, a summary of such model is presented. Early on, Chowdhury et al. [5] applied their method to publicly available chest X-ray images to identify COVID-19 based on CNN. To build their model, they extracted the crucial features from the input in a parallel manner. They achieved 96.58% prediction accuracy for three target classes (COVID-19 cases, healthy individuals, and the viral Pneumonia) on a dataset consisting of 2905. Later on, Sekeroglu et al. [6], implemented different machine learning models including, transfer learning, conventional models, and CNN to identify COVID-19 infected patients where the CNN model perform better with 98.50% prediction accuracy. However, they were unable to address the imbalance dataset issue. In a similar study, Elaziz et al. [7], proposed an ensemble of multi-CNN, Bayesnet and CFS to differentiate COVID-19 and non-COVID-19 images extracted from CT scanners. For feature extraction purpose, they used Fractional Multichannel Exponent Moments (FMEM), and Manta-Ray Foraging Optimization. At the same time, Hu et al. [8], proposed a system using supervised deep learning methods that maintain a weekly detection protocol in order to classify cases of COVID-19 efficiently. More recently, Sakib et al. [9], proposed a new method called DL-CRC using data augmentation strategy to enhance the detection performance up to 40% (from 54.55% to 93.94%). Later on, Elgendi et al. [10] proposed a DarkNet-19 deep learning model to tackle this problem with promising performance. At the same time, Heidari et al. [11], developed a CNN model to identify COVID-19 Cases from Chest X-ray Images. In this model the preprocessing performed by histogram and bilateral low pass filter and enhances the prediction performance to 94.5. Similarly, Minaee et al. [12], proposed a new CNN model and tested their model on over 3000 images to get a sensitivity rate of 98%. In another work by Nishio et al. a Deep neural network strategy has been used in which VGG16 was performed to construct the model. They also used a transfer learning method in which the first 10 layers were devoted to applying transfer learning with an accuracy level of over 83% [13]. After that Ismael et al. [14] used CNN model to extract features from the images of chest X-ray and obtained the most improved accuracy of 94.7% with Restnet50 and linear kernel SVM classifier. To resolve the scarcity of COVID-19 chest X-ray images, Loey et al. [15] gathered all available images of Covid patients and produced data sets using the Generative Adversarial Networks (GAN) process.

Furthermore, the detection technique is divided further, with the key component being the covid X-ray image, and the other images being included or omitted to verify the accuracy level. Later on, Jain et al. [16] proposed another CNN based deep learning method for the detection of COVID-19 from the chest X-ray images. To train this model, they used above six thousand image data and divided the dataset into training (above five thousand) and testing dataset (almost one thousand). This model produced a top accuracy level of 97.97%. In a similar study, Abraham et al., used a combination of multi-level CNN and CFS mechanism to determine out the features from the image dataset and also used Bayesnet for the classification purpose. They reported 97% accuracy for a dataset with 71 images. However, as the number of samples increased, their prediction performance rapidly dropped [17]. More recently, Goel et al. [18] proposed a new model called OptCoNet that was developed using CNN. The model was tested against different classification mechanisms, and the accuracy rate was improved to 97.78%. In another work [19], an integrative model was used to identify COVID-19 severity. In this model, an encoder is used to extract features, a decoder for segmentation, and a perceptron for classification. This approach achieved a ROC curve of more than 97% with a small dataset containing 1369 images. After that, in a separate study [20], authors implemented a four-stage deep learning model was proposed to identify and differentiate individuals with COVID-19 from healthy ones. The validation has been carried out using cross-validation of five folds and gained accuracy of about 98%. Recently, using a million images as a dataset EL et al., proposed a method for early detection of coronavirus disease by incorporating ResNet-101 CNN and gained 71.9% accuracy [21]. Most recently, Tougaccar et al. [22], reconstructed the data image by applying the Fuzzy Color mechanism for preparing step and used Support Vector Machine (SVM) as classification method and achieved 99.29% prediction accuracy.

## Proposed Methodology

The general architecture of our proposed method, CovidMulti-Net, is presented in Fig. 1. As shown in this figure, framework is based on transfer learning models for the early detection and classification of COVID-19 cases. A novel algorithm for the entire classification method is also presented in this section. The specifications about each phase of the “CovidMulti-Net” framework is discussed in the following subsections.

**Figure 1:**
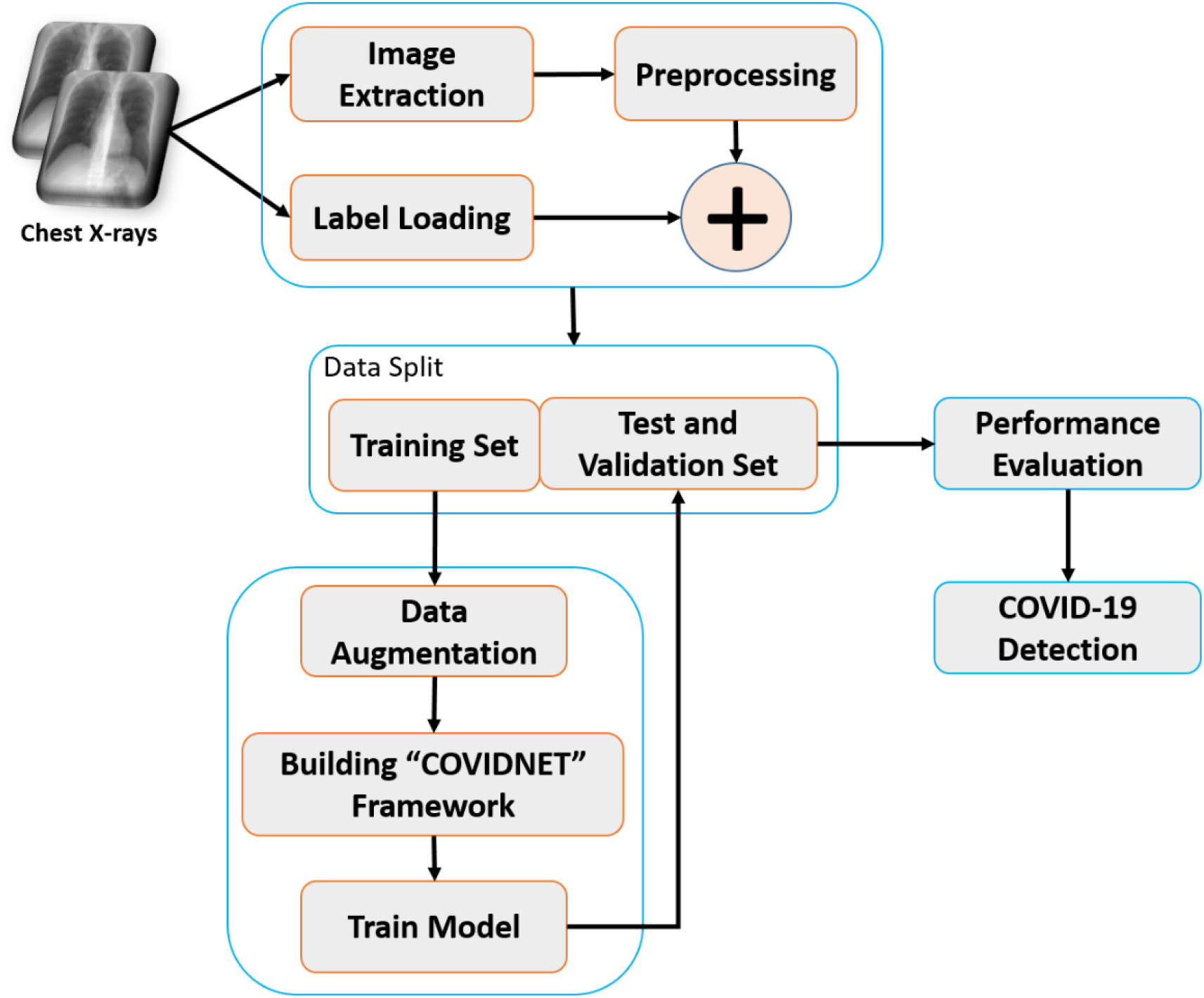
Proposed “CovidMulti-Net” framework for fast detection of COVID-19 cases.

### Data Collection

In this study, we utilize three datasets containing 2-Class (Normal and COVID-19), 3-Class (Normal, COVID-19, and Viral Pneumonia), and 4-Class (Normal, COVID-19, and Viral Pneumonia, Bacterial Pneumonia) CXR images.

### Class dataset

This dataset contains a total of 902 COVID-19 confirmed CXR images and 1341 normal CXR images. The COVID-19 CXR images were obtained from several publicly available image databases [23–27]. Those datasets contain both chest X-ray and CT images. We have excluded all the CT images and only select the CXR images with an anterior-posterior view for our work.

### 2-Class dataset

Furthermore, the second dataset is obtained from the COVID-19 Radiography Database, which was provided by the Kaggle community [27]. This dataset comprises 2905 CXR images, including 219 CXR images relating to the patient infected with COVID-19, 1341 normal images, and the other 1345 CXR images of patients with viral pneumonia.

### 3-Class dataset

The final dataset contains a total of 1639 CXR images where 320 CXR images related to the COVID-19 patients, 446 normal images, 424 CXR images of viral pneumonia patients, and 449 CXR images relating to the patients infected with bacterial pneumonia [28]. Fig 2 presents some samples of the images used in this study.

**Figure 2:**
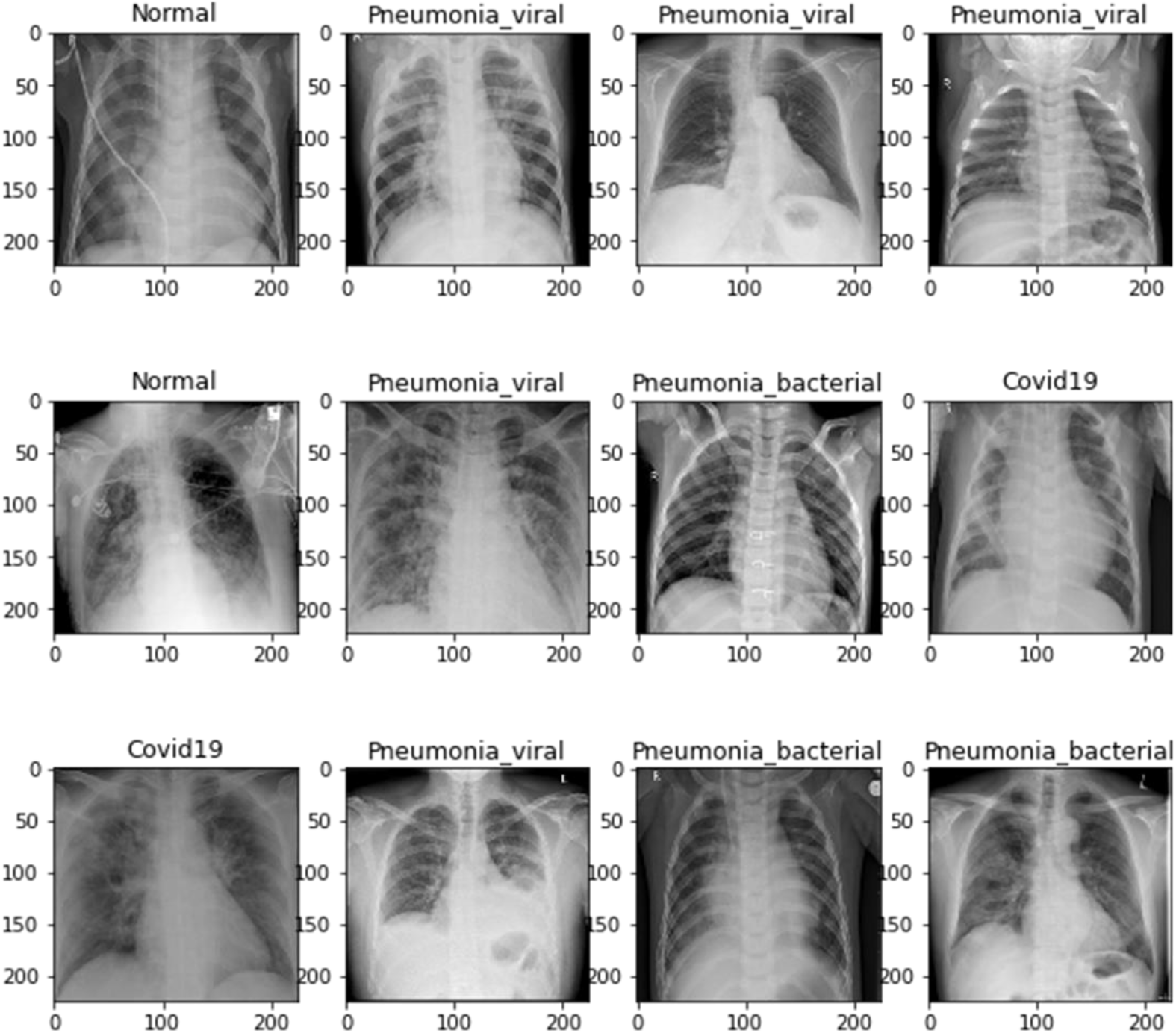
Sample of the images extracted from the dataset.

### Data-Preprocessing Stage

Before feeding the images into the proposed “CovidMulti-Net” framework, we preprocess all the images, including image resizing (224*224*3 pixels), format conversion (.png), and NumPy array conversion. The image resizing is performed as per the transfer learning concept, which help the proposed framework to achieve better performance in lower time and make the computation more straightforward. After that, we have shuffled all the image data before splitting them. In this way the “CovidMulti-Net” framework can train on unsorted data and stop relying on a narrow spectrum of the entire dataset. All the image datasets are then divided into three sub-sets, including training, validation, and testing sets, as described in Table 1. After that, a data augmentation technique is applied to the images. In this way, the “CovidMulti-Net” framework can define them as new ones. Besides, such a strategy also helps avoiding overfitting issues, expand the dataset size, and build a more generalized model [29]. We have performed various arbitrary photometric transformations with different parameter values as described in Table 2 in the 2-Class, 3-Class, and 4-Class datasets.

**Table 1:** Number of X-ray images used before data augmentation strategy.

| Dataset | Classification Type | Training | Validation | Testing |
| --- | --- | --- | --- | --- |
| 2-Class | Normal | 1001 | 250 | 90 |
|  | COVID-19 | 653 | 163 | 86 |
| 3-Class | Normal | 1001 | 250 | 90 |
|  | COVID-19 | 155 | 39 | 25 |
|  | Viral Pneumonia | 1000 | 250 | 95 |
| 4-Class | Normal | 294 | 74 | 78 |
|  | COVID-19 | 240 | 60 | 20 |
|  | Viral Pneumonia | 283 | 71 | 70 |
|  | Bacterial Pneumonia | 299 | 75 | 75 |

**Table 2:** Data augmentation approaches with different parameter value.

| Data Augmentation Strategy | Parameter Value |
| --- | --- |
| zoom_range | 2.5 |
| rotation_range | 90 |
| shear_range | 0.5 |
| width_shift_range | 0.5 |
| height_shift_range | 0.5 |
| horizontal_flip | True |
| vertical_flip | True |

As a result, the number of images is increased from 2243 to 14645, 2905 to 19075, and 1639 to 10015 after performing data augmentation strategy (see Table 3).

**Table 3:** Number of X-ray images used after data augmentation strategy.

| Dataset | Classification Type | Training | Validation | Testing |
| --- | --- | --- | --- | --- |
| 2-Class | Normal | 7007 | 1750 | 90 |
|  | COVID-19 | 4571 | 1141 | 86 |
| 3-Class | Normal | 7007 | 1750 | 90 |
|  | COVID-19 | 1085 | 273 | 25 |
|  | Viral Pneumonia | 7000 | 1750 | 95 |
| 4-Class | Normal | 2058 | 518 | 78 |
|  | COVID-19 | 1680 | 420 | 20 |
|  | Viral Pneumonia | 1981 | 497 | 70 |
|  | Bacterial Pneumonia | 2093 | 525 | 75 |

### CNN pre-trained models for feature extraction

After pre-processing the data, several CNN pre-trained architectures are adopted to separately extract the CXR images’ features. Next, we combined all the features using a concatenate layer for the classification task, as shown in Fig 3. In this work, we use three well-known up-to-date CNN pre-trained architectures, including DenseNet-169 [30], ResNet-50 [31], and VGGNet-19 [32], for the feature extraction task. The combined features represent high-level functionality such as sharpening, textures, roundness, and compactness of the CXR images [33]. After integrating all the extracted features, the proposed “CovidMulti-Net” framework tune 57 million training parameters that are greater than the existing DenseNet-169, ResNet-50, and VGG-19 architectures, respectively. Such high number of training parameters indicates a robust model for image recognition. The fundamental structure of each adopted CNN pre-trained architecture is outlined in the following sub-sections.

**Figure 3:**
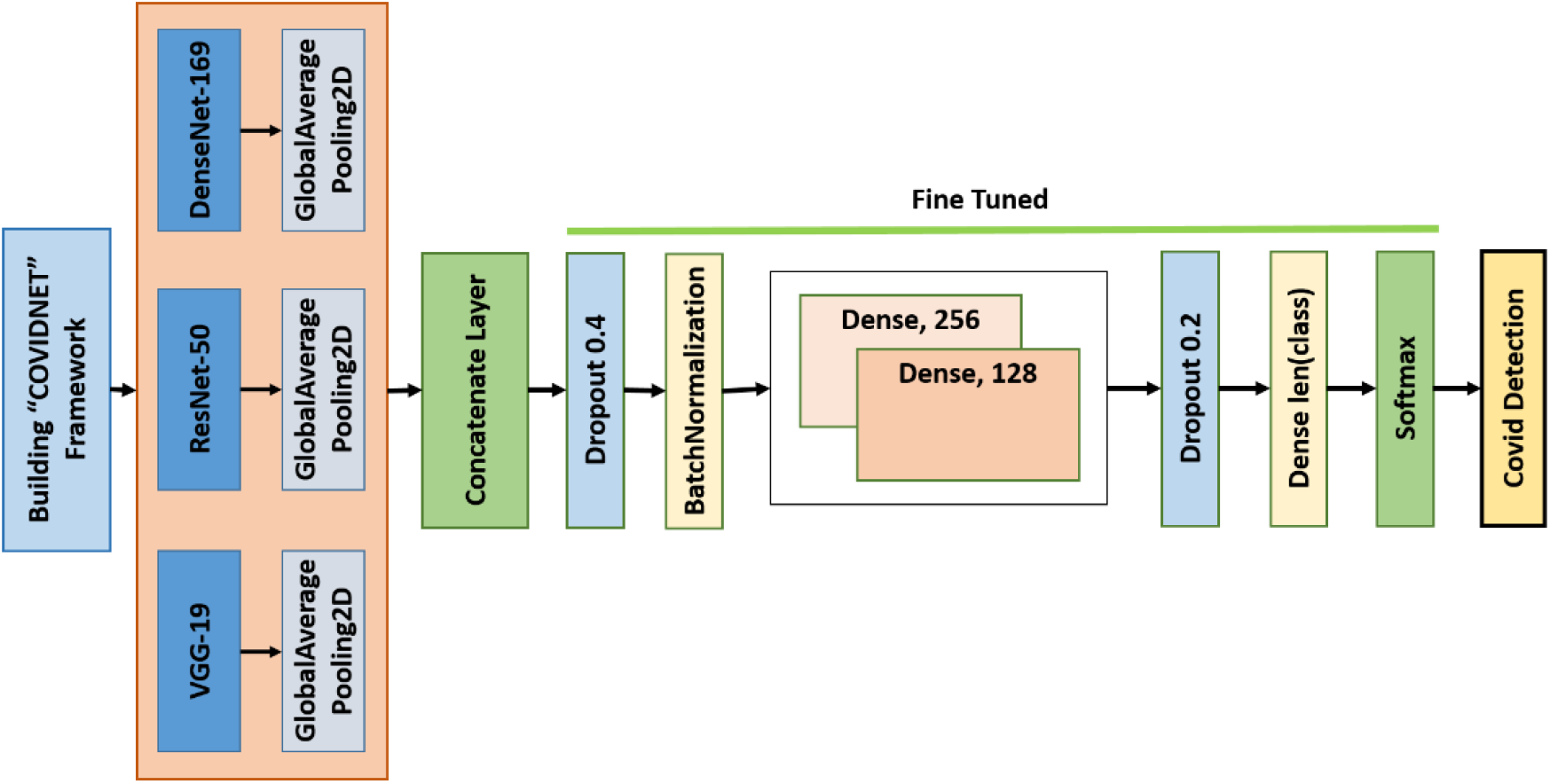
Fine-tuned process of the “CovidMulti-Net” framework

### DenseNet-169

DenseNet is considered one of the most advanced CNN architectures that demonstrates excellent classification accuracy on CIFAR-10, CIFAR-100, and ImageNet database in 2017 [30]. In this study, we utilize DenseNet-169 pre-trained architecture as our first feature extraction method, which contains 169 deep CNN layers. All the layers are densely connected to improve the declined accuracy caused by the vanishing gradient problem. Besides, the DenseNet-169 model provides more benefits in many ways, such as solving the overfitting issue, strengthening feature propagation, and reusing the model’s features.

### ResNet-50

He et al. popularized the concept of using deeper layers within a network by implementing the ResNet architecture that secured the first position at ILSVRC and COCO 2015 competition with a minimal error rate of 3.5% [31]. Here, we use ResNet-50 as our second feature extraction method, which consists of 50 convolution layers with five residual blocks. The residual blocks conserve the gained knowledge and speed up the training by boosting the model capacity. The ResNet pre-trained architecture also introduces shortcut connections that allow the data to flow easily between the layers and prevent information loss during training deep neural networks.

### VGG-19

Simonyan et al. first introduced VGGNet which become firs rank image localization and second position for image classification at the ILSVLC competition in 2014 [32]. VGGNet performs better than the AlexNet architecture because of its simple structure. In this work, we have adopted VGG19 as our final feature extraction method which contains 16 convolution layers with three fully connected layers.

### Fine-tuning Process

In Fig. 3, the architecture of our model is shown as a reflection of our fine-tuned model. After computing the features using three pre-trained models mentioned above, a concatenate layer merges all the features. Then the features were passed through other additional layers such as dropout, batch normalization, and dense layers. The last dense layer outputs decisions as the number of classes remain in the input data. Following a softmax layer, the final decision is made for particular input features. The proposed “CovidMulti-Net” framework for detecting COVID-19 cases is given as Algorithm 1.

## Experimental Setup, Performance Metrics and Results Analysis

This section illustrates the experimental configuration and the findings obtained using the proposed “CovidMulti-Net” framework on three datasets. To evaluate the proposed framework’s efficiency, we have used statistical parameters like Precision, Recall, F1-Score, False-positive rate, and True negative rate.

### Experimental Setup

Here we used Keras, as an open-source library that interfaces python with the neural network to build “CovidMulti-Net”. We also use Google Co-lab, a free online cloud service that allows the use of Tesla K80 GPU with 12 GB of GDDR5 RAM, Intel Xeon processors with two @2.2 GHz cores, and a total of 13 GB ram, to conduct all the computations.

#### Algorithm 1 Automated COVID-19 detection and classification from CXR images.

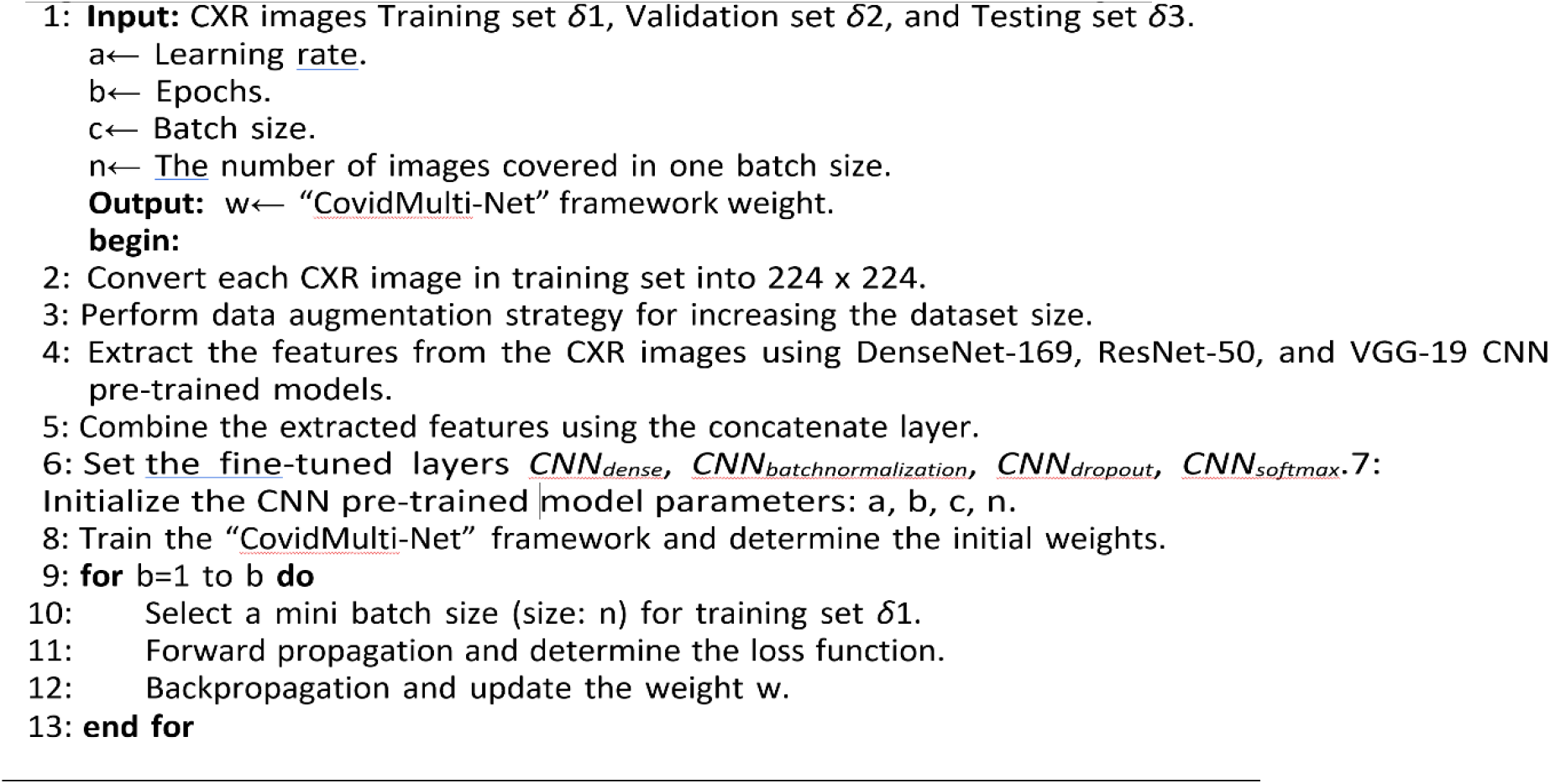

### Performance evaluation metrics

To evaluate the performance of the proposed framework, six statistical parameters including accuracy (Acc), precision (Pre), recall (Rec), false positive rate (FPR), true negative rate (TNR), and F1-Score are used. The performance metrics are:

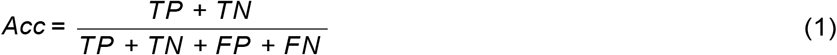

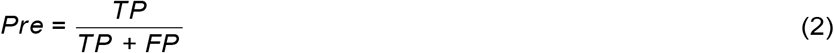

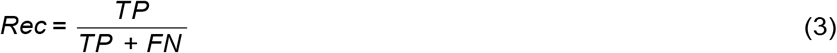

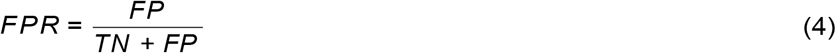

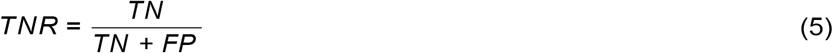

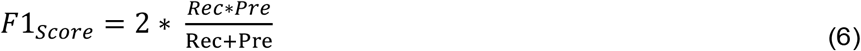

Herein, TP and TN refer to the correctly predicted positive and negative covid cases, whereas FP and FN correspond to the number of incorrectly predicted positive and negative covid cases. In addition, we also compute the area under curve (AUC) of the proposed “CovidMulti-Net” framework. This curve helps to identify the overall achievement of the “CovidMulti-Net” framework and also reveals a trade-off between precision and recall.

### Training and Parameter Optimization

A deep neural network is specifically simulated in this paper for the identification of binary and multiclass COVID-19 instances. In Table 4, all the hyper-parameters tuned in this study with their values are listed. One of the main parameters of the model is the optimizer function. For that, we used Adam optimizer function to tune for both binary and multiclass datasets. The key properties of the AdaGrad and RMSProp optimizers can be combined with this function. Furthermore, we also use binary cross-entropy as a loss function to train the binary class dataset. On the other hand, for multiclass datasets, we use categorical cross-entropy. To reduce the loss function, an optimal value for the learning rate is required. However, it is a difficult process to determine the optimal learning rate. Here we set the learning rate to 0.0001 as it has been shown as an effective number for deep learning classification.

**Table 4:**
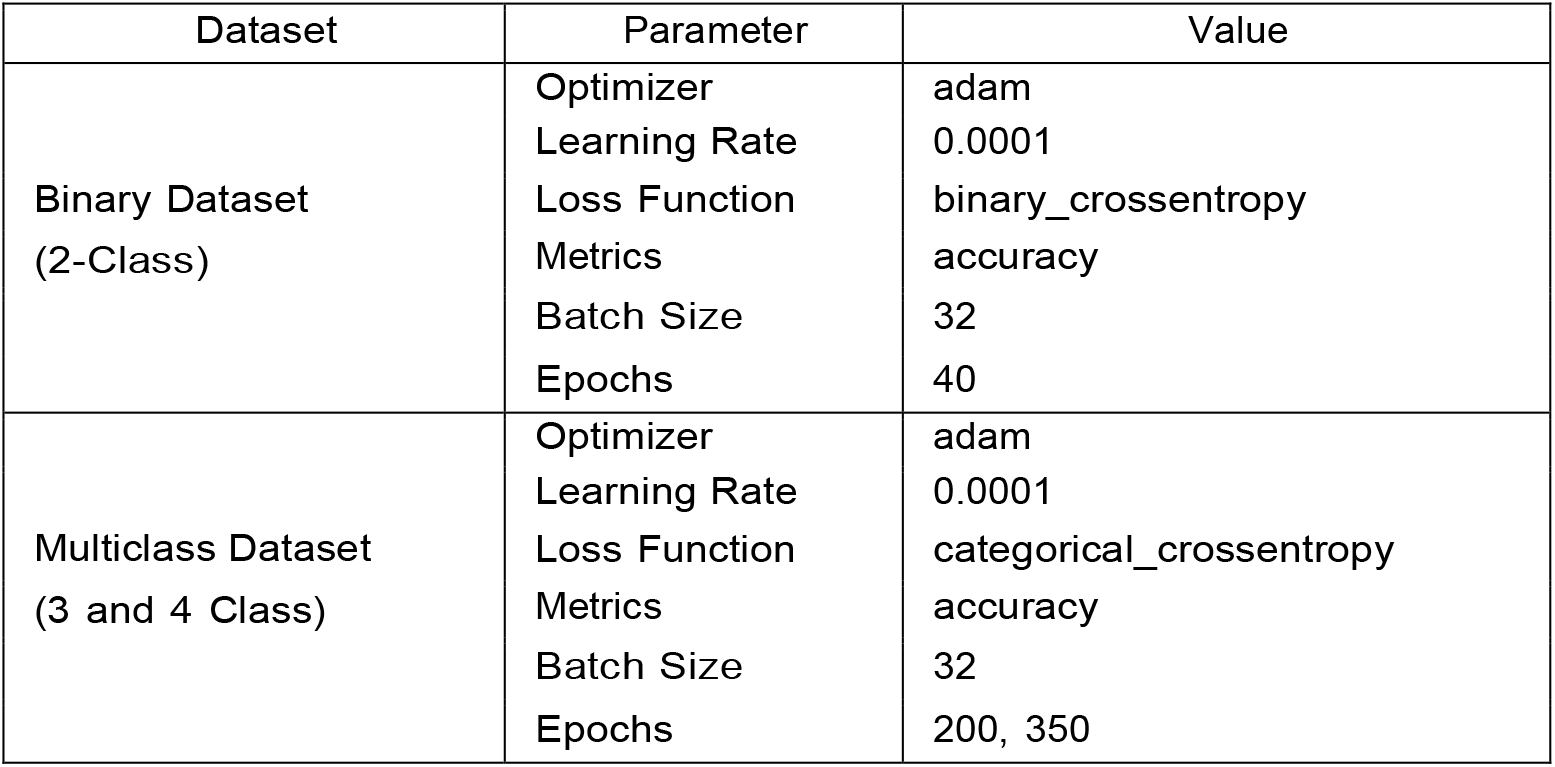
The effective parameters used in the “CovidMulti-Net” framework

Fig. 4, Fig. 5 and Fig. 6 reflect the simulation results of training the proposed “CovidMulti-Net” framework for different types of datasets. From the results, it is observed that the problem of overfitting is not present during model training since the results are consistent on all the employed datasets. As shown in Fig. 4(a), the 2-class dataset is trained for 40 epochs and the model managed to achieve more than 97% training accuracy and 99.0% validation accuracy after only 35 epochs of training. In Fig. 4(b), the curve is starting to dramatically reduce the loss value. However, due to the small batch size of 32, some fluctuations have arisen. Fig. 5(a) illustrates the performance of the proposed model for the 3-class dataset that needed to be further trained. The framework is trained for 200 epochs, and 94% training accuracy has been achieved right after the 180th epoch. Also, the average accuracy of the validation stage is 95.4%. The loss function curve is depicted in Fig. 5(b) where the loss value is almost zero, and during the training and validation, fewer fluctuations occurred. After that, we trained the “CovidMulti-Net” framework on a 4-class dataset for 350 epochs, and the model achieved more than 93.0% training accuracy and 95.0% validation accuracy depicted in Fig. 6(a). Fig.6(b) represents that the magnitude of the loss is simply reduced to 0.4.

**Figure 4:**
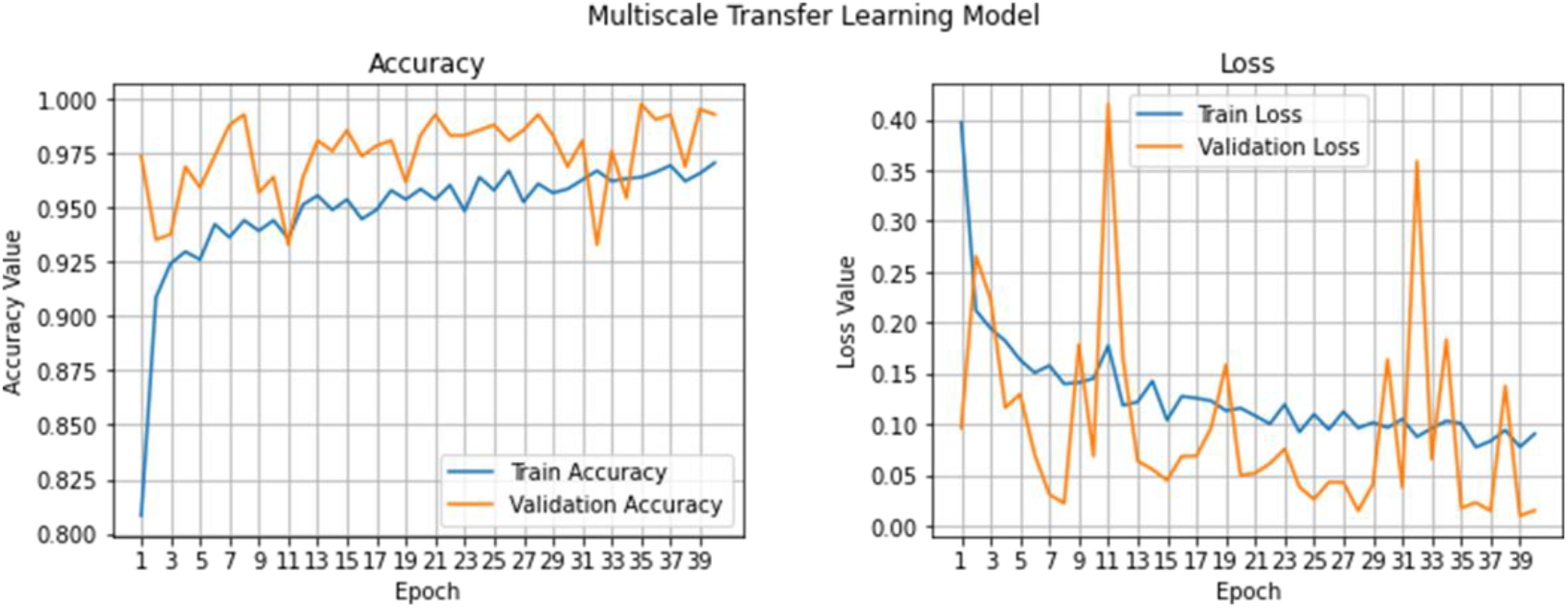
Training progress for 2-class dataset: (Left) training and validation accuracy (higher is better), and (Right) training and validation loss (lower is better)

**Figure 5:**
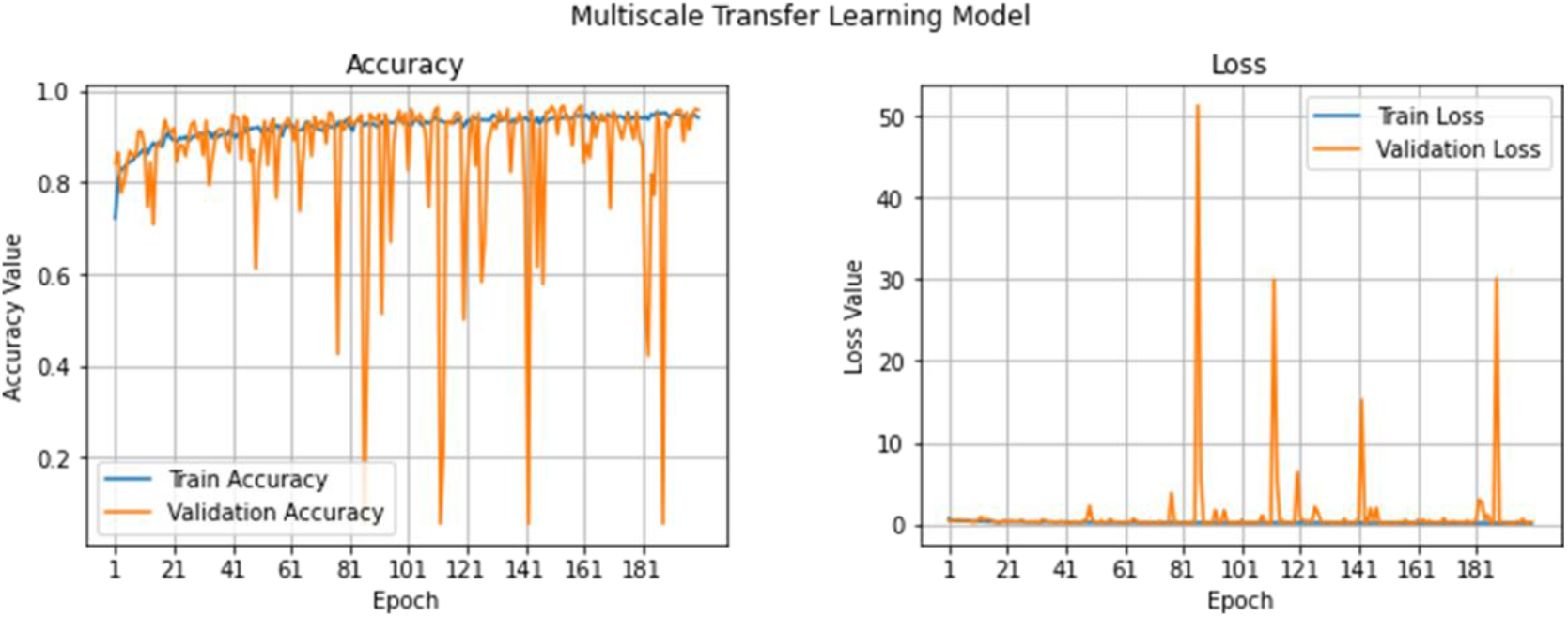
Training progress for 3-class dataset: (Left) training and validation accuracy (higher is better), and (Right) training and validation loss (lower is better)

**Figure 6:**
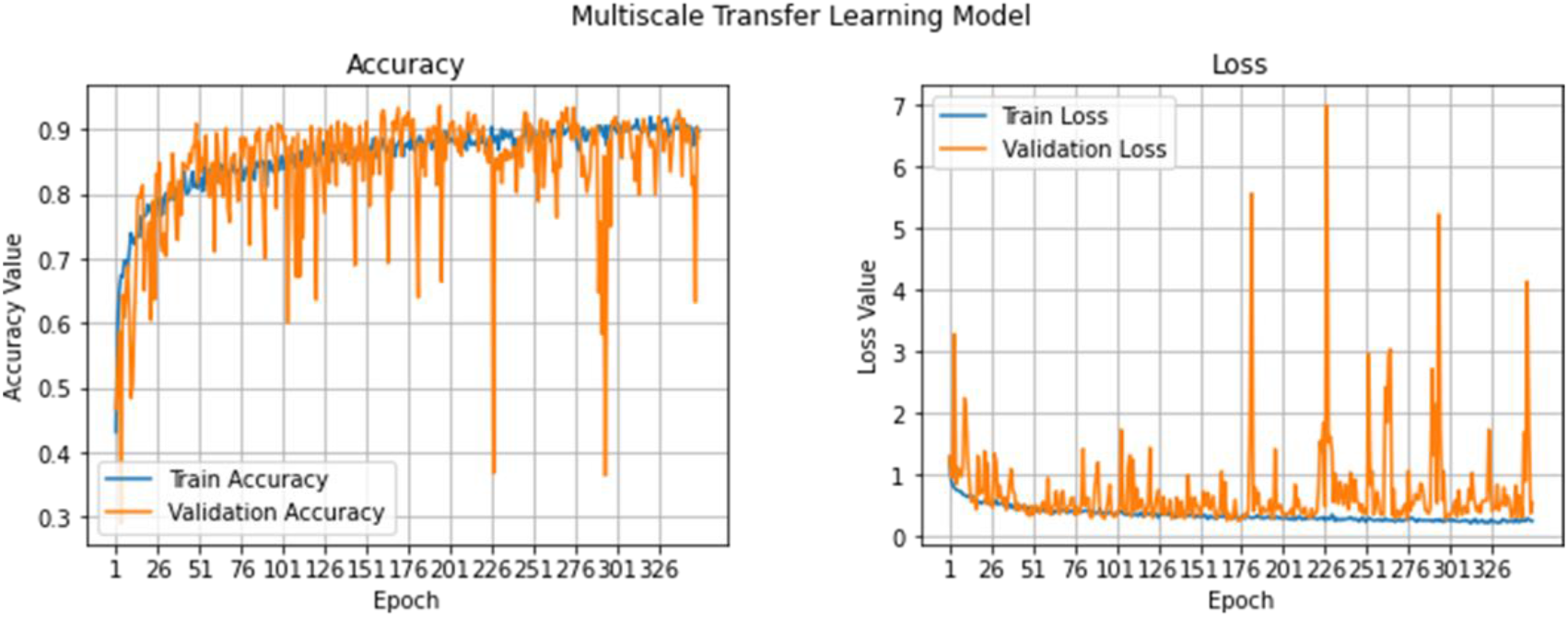
Training progress for 4-class dataset: (Left) training and validation accuracy (higher is better), and (Right) training and validation loss (lower is better)

### Result Analysis

Fig. 7 presents the confusion matrix and the ROC curve for the 2-class dataset.It is obvious from Fig. 7(Left) that a total of 89 and 86 CXR images of nor-mal and COVID-19 cases are perfectly classified. At the same time, the proposed “CovidMulti-Net” framework misclassifies only one normal CXR image. We have also obtained an area value of 0.994 (see Fig. 7(Right)), which shows the stability of the “CovidMulti-Net” framework in classifying both positive and negative classes.

**Figure 7:**
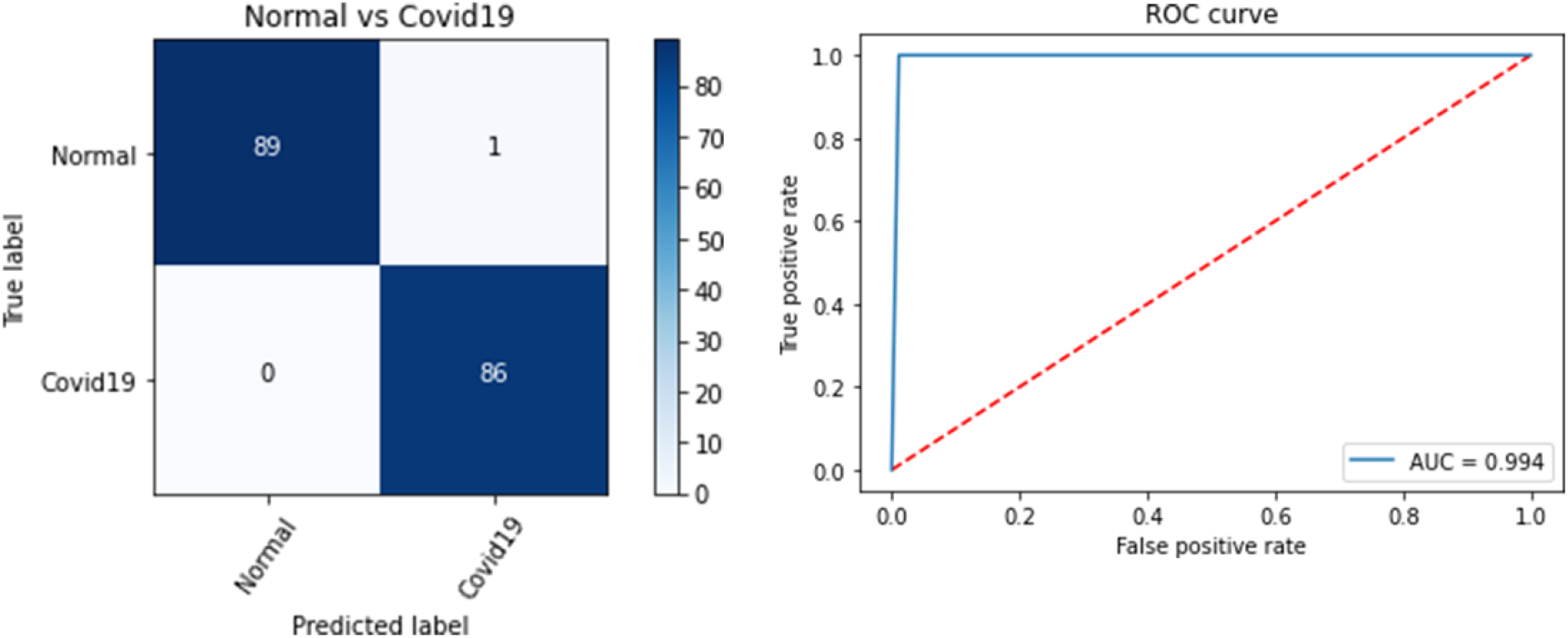
“CovidMulti-Net” performance on 2-Class dataset. (Left: confusion matrix, Right: ROC curve.)

Furthermore, from Fig. 8(Left) it can be observed that a total of 25, 91 and 79 CXR images are properly classified for the COVID-19, pneumonia and normal cases. However, the proposed “CovidMulti-Net” framework failed to identify 11 normal and 4 CXR images infected with pneumonia virus. Here, the area under the curve value is 0.931 (see Fig. 7(Right)) for the 3-class dataset.

**Figure 8:**
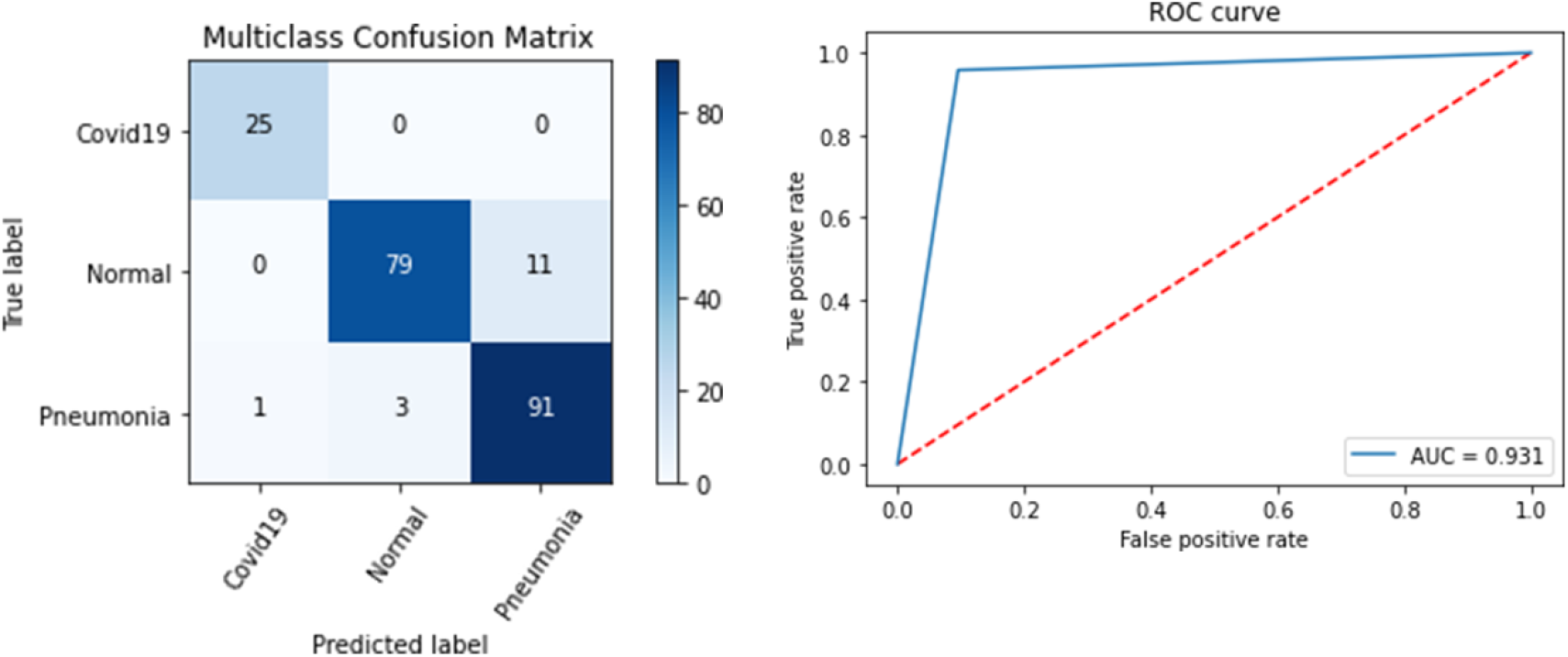
“CovidMulti-Net” performance on 3-Class dataset. (Left: confusion matrix, Right: ROC curve)

Finally, from Fig. 9(Left) it can be observed that a total of 20 CXR images are correctly identified as COVID-19, 56 as viral pneumonia, 67 as bacterial pneumonia, and 83 as normal X-ray images. Here, the area under the curve is 0.928 as shown in Fig. 9(Right). The most notable advantage of “CovidMulti-Net” framework is that it does not misclassify any CXR image affected by COVID-19 for all these three datasets.

**Figure 9:**
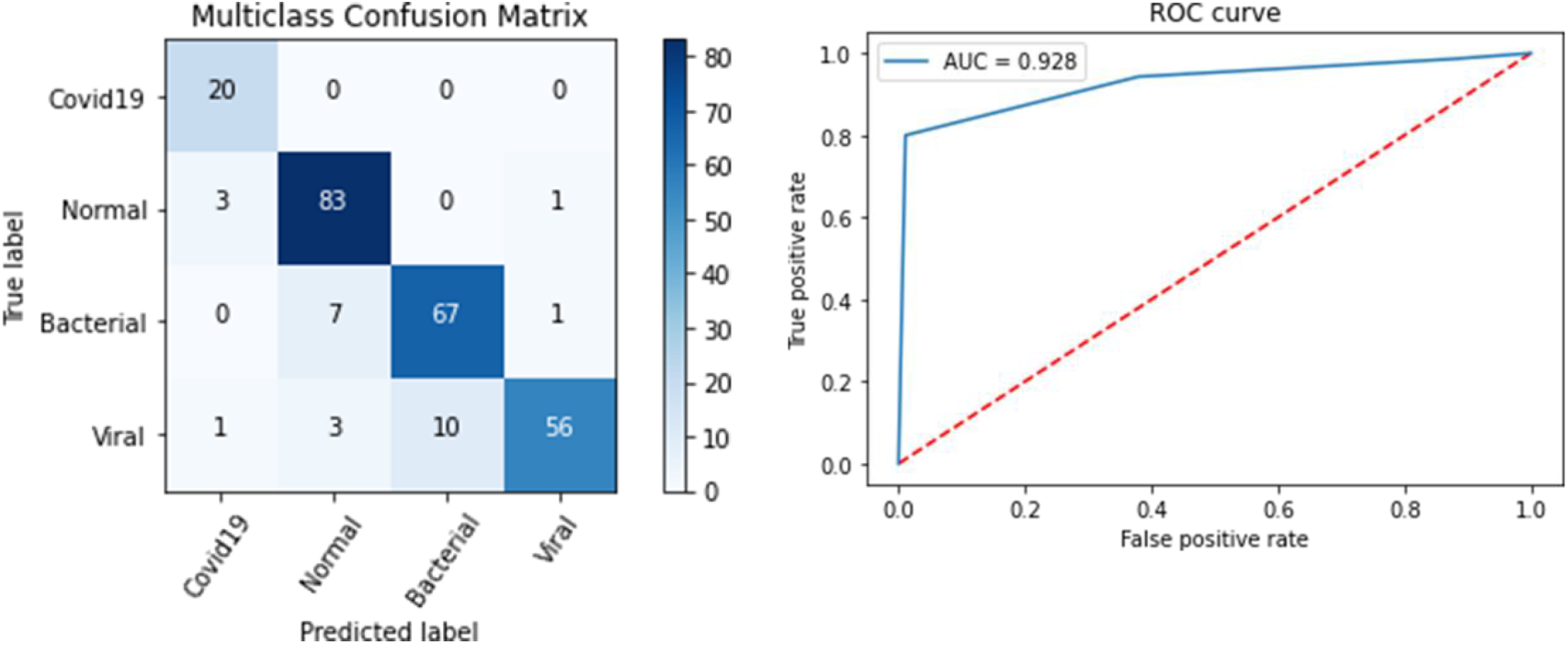
“CovidMulti-Net” performance on 4-Class dataset. (Left: confusion matrix, Right: ROC curve)

The comparative analysis among the three datasets using the proposed classifier is presented in Table 5. This table demonstrates that the highest classification accuracy is achieved when the “CovidMulti-Net” framework trained on the binary dataset. The average accuracy is 99.4% for this 2-class classifier. However, when the same model was retrained as a tertiary and multiclass classifier, the mean accuracy decreased a little bit, though the numerical value remains over 94% in both of these cases. While the number of output classes started to increase, the machine learning model needs to generalize the features more accurately so that it can key out the target outputs. Moreover, the larger number of classes requires a larger number of samples to train and validate the model. Hence, it is possible to conclude why the model resulted in a lower accuracy as the number of classes is increased. Apart from the classification accuracy, some other performance metrics results are also listed in Table 5.

**Table 5:** Results obtained using the “CovidMulti-Net” architecture on three publicly accessible datasets.

| Data set | Category | T<br>P | T<br>N | F<br>P | F<br>N | Accur<br>acy | Precisi<br>on | Reca<br>ll | FPR | TNR | F1-score |
| --- | --- | --- | --- | --- | --- | --- | --- | --- | --- | --- | --- |
| 2-Clas<br>s | Normal | 89 | 86 | 0 | 1 | 99.4 | 1.0 | 0.99 | 0 | 1.0 | 0.99 |
|  | COVID-19 | 86 | 89 | 1 | 0 | 99.4 | 0.99 | 1.0 | 0.01 | 0.99 | 0.99 |
|  | Average Score |  |  |  |  | 99.4 | 0.99 | 0.99 | 0.005 | 0.99 | 0.99 |
| 3-Clas<br>s | Normal | 79 | 11<br>7 | 3 | 11 | 93.3 | 0.96 | 0.88 | 0.025 | 0.97 | 0.92 |
|  | COVID-19 | 25 | 18<br>4 | 1 | 0 | 99.5 | 0.96 | 1.0 | 0.005 | 0.99 | 0.98 |
|  | Viral<br>Pneumonia | 91 | 10<br>4 | 11 | 4 | 92.8 | 0.89 | 0.96 | 0.09 | 0.91 | 0.92 |
|  | Average Score |  |  |  |  | 95.2 | 0.94 | 0.95 | 0.04 | 0.96 | 0.94 |
| 4-Clas<br>s | Normal | 83 | 15<br>5 | 10 | 4 | 94.4 | 0.89 | 0.95 | 0.06 | 0.94 | 0.92 |
|  | COVID-19 | 20 | 22<br>8 | 4 | 0 | 98.4 | 0.83 | 1.0 | 0.02 | 0.98 | 0.91 |
|  | Viral<br>Pneumonia | 56 | 18<br>0 | 2 | 14 | 93.6 | 0.97 | 0.80 | 0.01 | 0.99 | 0.88 |
|  | Bacterial<br>Pneumo-<br>nia | 67 | 16<br>7 | 10 | 8 | 92.8 | 0.87 | 0.89 | 0.06 | 0.94 | 0.88 |
|  | Average Score |  |  |  |  | 94.8 | 0.89 | 0.91 | 0.04 | 0.96 | 0.90 |

## Discussion

Many COVID-19 classification studies have provided versatile architectures that incorporate several issues. Compared to the existing literature with various frame-works, parameters, and depths, the performance of the “CovidMulti-Net” frame-work has been summarized in Table 6. In most cases, ResNet model has been used, which outperforms the VGG styled CNN pre-trained model but it is not much effective when dealing with a large number of images [11] [34–36]. It is apparent from Table 6 that the proposed “CovidMulti-Net” framework shows the best prediction accuracy in identifying both binary an multi-class CXR images compared to the prior work. By properly integrating all the pre-trained models, we have obtained an average prediction accuracy of 99.4% for 2-class, 95.2% for 3-class and 94.8% for 4-class datasets, respectively. Apart from that, the proposed “CovidMulti-Net” framework provides a segmentation-free feature extraction technique that does not involve any handcrafted features. Besides, the COVID-19 affected CXR images were misclassified by all the methods found in the literature, which is more dangerous and decreases the chances of survival [11] [28] [34–41]. Our proposed method, however, does not misclassify any images affected by COVID-19 in both binary and multiclass datasets. Fig. 10 depicts some of the CXR images that the “CovidMulti-Net” framework predicts. The bio-medical datasets are going to be bigger and bigger. In the analysis of such data, application of machine learning and deep learning techniques has become more attractive given the rising complexity of the data [42-46]. Therefore, it is important to implement novel techniques to uncover the biomedical patterns, in particular biomedical imaging data. The proposed deep learning model has a great potential to be used on healthcare imaging data analysis including CXR images, brain imaging, etc.

**Table 6:** Performance Comparison of the proposed “CovidMulti-Net” framework with other methods in binary and multiclass classification.

| Study | Image type | Accuracy (2-Class) | Accuracy (3-Class) | Accuracy (4-Class) | Method |
| --- | --- | --- | --- | --- | --- |
| Hemdan et al. [34] | X-ray | 90.0 | - | - | VGG19, DenseNet121 |
| Narin et al. [35] | X-ray | 98.0 | - | - | ResNet50, ResNetV2, InceptionV3 |
| Sethy et al. [36] | X-ray | 95.4 | - | - | ResNet50+SVM |
| Heidari et al. [11] | X-ray | 98.1 | 94.5 | - | VGG16 |
| Tulin et al. [37] | X-ray | 98.08 | 87.02 | - | DarkNet |
| Xu et al. [38] | CT | - | 86.7 | - | ResNet |
| Toraman et al. [39] | X-ray | 97.24 | 84.22 | - | CapsNet |
| Khan et al. [28] | X-ray | 98.8 | 94.5 | 89.6 | Xception |
| Wang et al. [40] | X-ray |  | 93.3 | - | CovidNet |
| Apostolopoulos et al. [41] | X-ray | 96.78 | 94.72 | - | MobileNetV2 |
| Proposed Method | X-ray | 99.4 | 95.2 | 94.8 | DenseNet-169 + ResNet-50 + VGG-19 + Fine-tuned |

**Figure 10:**
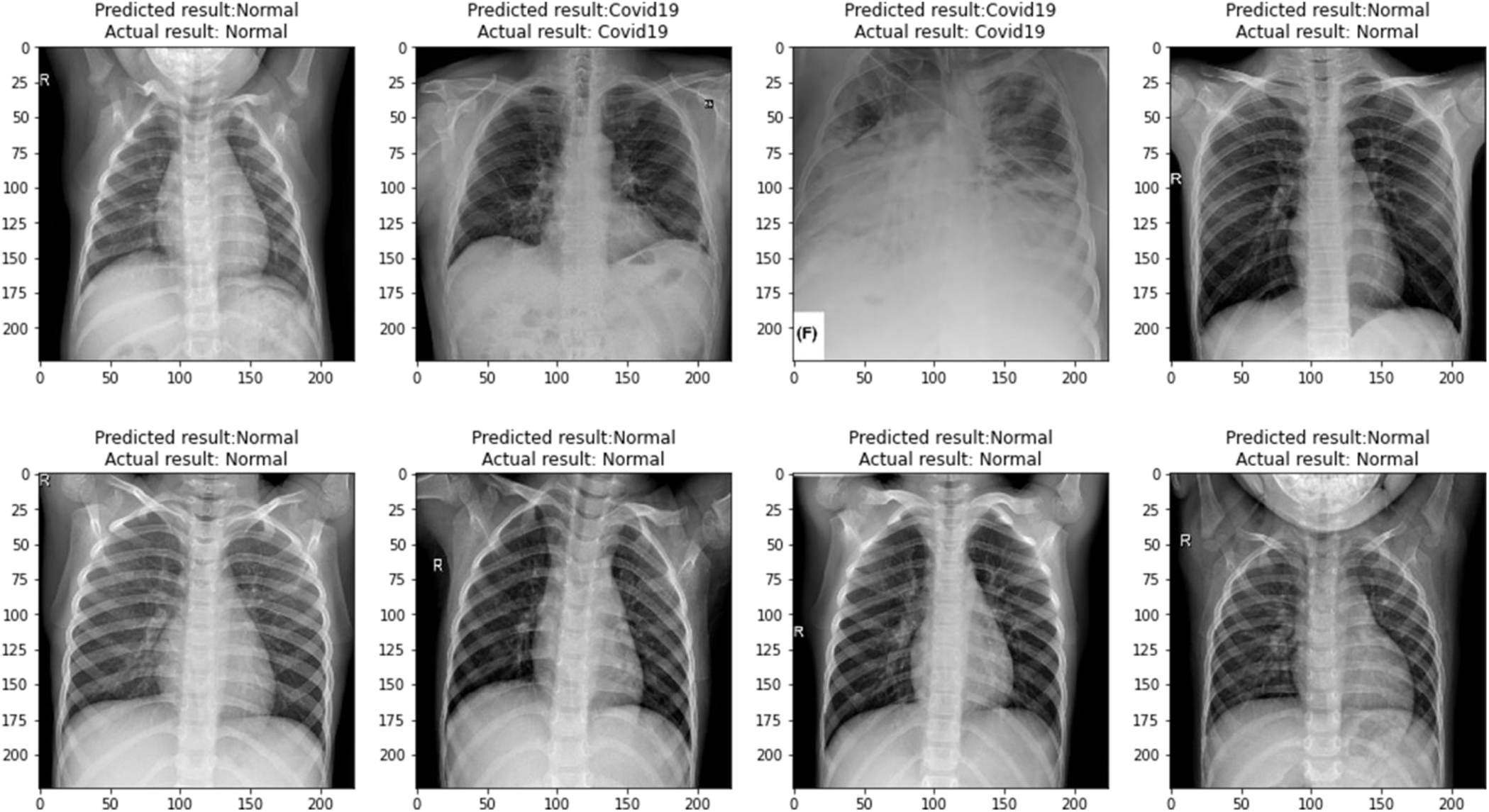
“CovidMulti-Net” evaluated some X-ray images.

## Conclusion

Since COVID-19 pandemic poses a severe threat to public health, it is essential to identify all the positive cases and treat them as soon as possible. With this in mind, this research introduces a “CovidMulti-Net” framework based on deep transfer learning concept to detect COVID-19 cases from chest X-ray images. This framework consists of three CNN pre-trained models, including DenseNet-169, ResNet-50, and VGG-19, for the feature extraction. After that, all the extracted features are combined using a concatenate layer. The combined features are then fine-tuned using three fully connected layers for the classification task. We evaluated the “CovidMulti-Net” framework on three publicly available datasets. Experimental results indicate that we have achieved 99.4%, 95.2%, and 94.8% classification accuracy for 2-class, 3-class, and 4-class datasets, respectively. Besides, the proposed approach is computationally inexpensive and achieved the highest classification accuracy compared to the other state -of-the-art models. However, the accuracy may be further improved once more training data becomes available. Also, the proposed approach still needs clinical study and testing, but with such high performance, we believe that the “CovidMulti-Net” framework would be an outstanding candidate for COVID-19 detection.

## Data Availability

All the data are publicly available as mentioned in the manuscript.

## Availability of data and materials

All the materials are available at https://github.com/saikat15010/CovidMulti-Net-Architecture.git.

## Authors’ contributions

SS designed the study. SIK, AR, and MRK wrote the manuscript; AR, and NIB collected data. SS, AR, AD and HAR edited the manuscript; SIK, AR, and MRK carried out the analyses. SIK, and AR generated all figures and tables. HAR, was not involved in any analysis. HAR, AD and SS do not have any responsibility against scripts, analyses and figures All authors have read and approved the final version of the paper.

